# A systematic review of the effects of traditional East Asian medicine on symptom clusters during the menopausal transition

**DOI:** 10.1101/2022.04.26.22274224

**Authors:** Lisa J. Taylor-Swanson, Athena Sdrales, Rana Ali, Belinda Anderson, Lisa Conboy, Melissa Cortez, Xiaoming Sheng, Cynthia Price, Nancy Fugate Woods, Julie Fritz, Paula Gardiner

## Abstract

**Importance:** Given that many midlife women use evidence-based non-pharmacologic interventions for symptom management during the menopausal transition and early postmenopause and that many women experience two or more symptoms (symptom cluster), it is important to review recent evidence on said interventions for symptom clusters. This review focuses on randomized clinical trials (RCTs) of traditional East Asian Medicine (TEAM), including acupuncture, acupressure, moxibustion, and Chinese herbal medicine, for hot flashes and one or more co-occurring symptoms experienced during peri- or post-menopause.

**Objective:** The primary objective of the study was to review RCTs (published 2011-2021) of TEAM interventions for hot flashes and at least one other co-occurring symptom, including sleep problems, cognitive function, mood, and pain.

**Evidence Review:** We searched Medline, CINAHL Plus, and PsychINFO for RCTs reported in English from July 7, 2011, to December 31, 2021. We included RCTs that assessed women in the menopausal transition or early postmenopause with masking as appropriate; TEAM interventions were studied as the treatment with one or more comparison groups; hot flushes and at least one additional symptom from at least one of the symptom clusters were reported as an outcome (from sleep, mood, cognition, pain symptom groups). Bias was assessed.

**Findings:** Of 409 abstracts identified, 9 RCTs examined the effectiveness of therapies for hot flashes and at least one additional co-occurring symptom. One study reported separately on two TEAM interventions. The maximum trial duration was 6 months (range: 5 weeks – 6 months). Statistically significant improvement was reported in 2 or more symptoms: 5 of 6 studies of acupuncture, one acupressure study, one moxibustion study, 1 of 2 studies of Chinese herbal medicine.

**Conclusions and Relevance:** Our systematic review summarizes the recent literature on TEAM interventions for symptom clusters during the MT and EPM. A majority of studies reported symptom reduction. Overall, our findings highlight the need for further investigation with studies that include whole systems TEAM as each intervention was studied separately in the reviewed trials. Clinical practice often includes concurrent use of acupuncture, moxibustion, CHM, and advice to patients to use self-acupressure. Studying each modality separately is a scientific abstraction that does not reflect clinical practice.

**Key Points:** *Question/Objective:* What is the effect of traditional East Asian medicine (TEAM) therapeutics, including acupuncture, acupressure, Chinese herbal medicine, or moxibustion for hot flashes and one or more co-occurring symptoms during the menopausal transition (MT)?

*Findings:* Nine RCTs (n=811) evaluating the effects of TEAM therapeutics on MT symptoms were included. Statistically significant improvement was reported in these studies: acupuncture (5 of 6), 1 acupressure study, 1 moxibustion study, and Chinese herbal medicine (1 of 2).

*Meaning:* Based on the results of this systematic review, TEAM interventions demonstrate promising improvement of hot flashes and co-occurring symptoms experienced during the MT.

## Introduction

The menopausal transition (MT) is a normal transition experienced by all individuals with a uterus^1^. Approximately 80% of women in the US experience symptoms during the MT and early postmenopause (EPM). Common symptoms include hot flashes and night sweats (vasomotor symptoms), pain of various types (musculoskeletal, arthralgias), awakening at night and trouble falling asleep, problems with mood such as anxiety and depression, and cognitive concerns such as difficulty concentrating and forgetfulness^1^. Some women experience more than one symptom concurrently, known as a symptom cluster. Symptom clusters are defined as multiple co-occurring symptoms that may or may not share etiology and vary with the menopausal transition and postmenopause stages. These clusters include hot flushes, mood, sleep, cognitive, and pain symptoms of varying intensity^2-5^.

Current first-line treatments are hormone therapies (HT)^6,7^ which have demonstrated efficacy for the treatment of vasomotor symptoms (hot flashes, sweats) and vaginal dryness and play a role in osteoporosis prevention for some women^8^. Yet, among perimenopausal (PMP) women surveyed about HT, 65% of those familiar with HT would not consider taking it^9^. Thus, some women utilize non-pharmacological interventions for symptom management. For example, a 2013 review of 9 surveys of perimenopausal and postmenopausal women indicated that 50.5% of women used complementary and integrative medicine for symptom management^10^.

Given that women in the MT and EPM are utilizing non-pharmacological interventions for symptom management, reviewing extant evidence on said interventions is important. This review is focused on traditional East Asian medicine (TEAM) interventions, including acupuncture, acupressure, moxibustion, and Chinese herbal medicine. The review is of TEAM interventions and their effect on MT and EPM symptoms as evidenced in randomized controlled clinical trials (RCTs). We included hot flashes and one or more symptoms (a symptom cluster) as this is what women typically experience and because the focus of multiple symptoms aligns with TEAM theory. TEAM differential diagnoses vary by clusters of presenting symptoms: TEAM clinicians, Licensed Acupuncturists, ask patients about their presenting symptoms across systems to formulate the TEAM differential diagnosis. The study objective is to systematically review the effects of TEAM therapeutics on hot flashes and co-occurring symptoms in women experiencing a natural menopausal transition. All but one^11^ prior systematic reviews of TEAM therapeutics for symptoms during the menopausal transition focus on one primary symptom and typically focus on one primary intervention, as well. This review aims to fill this gap by identifying the reported therapeutic effects of TEAM interventions, including acupuncture, acupressure, Chinese herbal medicine (CHM), and moxibustion on symptom clusters comprised of hot flashes co-occurring symptoms, including pain, sleep, mood, and cognitive concerns.

## Methods/Literature Search

This systematic review was performed following Preferred Reporting Items for Systematic Reviews and Meta-Analyses (PRISMA) guidelines^12^. A PubMed/Medline (National Library of Medicine), CINAHL Plus (EBSCO), and PsychINFO search of the primary literature was conducted on January 5, 2022. The search included variant spellings and truncations of the following terms – menopause, climacteric, premenopause, perimenopause, postmenopause, hot flashes – along with various combinations of acupuncture, traditional Chinese medicine, traditional East Asian medicine, acupressure, moxibustion, and Chinese herbal medicine terminology. Retrieval was limited to publications in the English language, human, and published July 7, 2011 through December 31, 2021. The review was registered in the International Prospective Register of Systematic Reviews (PROSPERO 307590) and did not require institutional review board approval. We included acupuncture, acupressure, Chinese herbal medicine, and moxibustion. We excluded literature reviews, systematic reviews, and studies that did not include a TEAM intervention. All retrieved citations were screened in duplicate (LJTS and AS or LJTS and RA). Articles that met inclusion criteria were manually searched for additional articles.

This study focused on published randomized trials of hot flashes and one or more co-occurring symptoms, called a symptom cluster, including hot flashes, pain, sleep, mood, and cognitive concerns. The rationale is that this symptom cluster is identified in the scientific literature on the menopausal transition^2-5^ and that clusters of symptoms are likewise examined to determine the traditional East Asian medicine (TEAM) differential diagnosis. Yet, while the importance of symptom clusters has been identified scientifically and clinically, there are no models yet in the scientific literature for examining symptom clusters as outcomes. We operationalized a symptom cluster for this project: when two or more symptoms (including hot flashes, pain, sleep, mood, and cognitive concerns) were measured in the same study as primary or secondary outcomes. Symptom groups and their representative symptoms consist of hot flashes (hot flashes, hot flushes, vasomotor symptoms), pain (joint pain, joint ache, musculoskeletal pain, backache, headache, arthralgias), sleep concerns (awakening at night, early morning awakening, difficulty getting to sleep, or sleep disorders), mood (depressed mood, mood changes, crying, irritability, melancholia or anxiety), and cognitive concerns (difficulty concentrating, forgetfulness, poor memory).

### Study selection

Two authors independently screened all citations from the databases searches (LJTS and AS or LJTS and RA). Disagreements were resolved through discussion. We erred on the side of inclusion in full-text screening, which the same two reviewers performed.

### Inclusion and exclusion criteria

Inclusion criteria included: randomized trials in which women in the menopausal transition or postmenopause participated; masking was used as appropriate; non-pharmacologic therapies were studied as the treatment with one or more comparison groups; hot flushes and at least one additional symptom from at least one of the symptom clusters was reported as an outcome (from sleep, mood, cognition, pain symptom groups). We excluded papers not published in English and samples that included cancer, HIV, or other concurrent diagnoses.

### Data collection and synthesis procedures

One author performed data extraction (either AS or RA), which was then verified by one reviewer (LJTS). Discrepancies were resolved through discussion and referring back to the article. The data extraction form included fields about the article (author, year, country), design (randomization, number of groups, masking), main intervention and comparison or control, outcomes (hot flashes, other symptoms), and results and adverse events (hot flashes, other symptoms) (see Appendix 1). Cochrane levels of evidence ratings were noted, and the risk of bias (RoB) in RCTs was assessed using the Cochrane RoB2.^13^ The domains included in RoB 2 cover all types of bias that are currently understood to affect the results of randomized trials. These are: bias arising from the randomization process; bias due to deviations from intended interventions; bias due to missing outcome data; bias in measurement of the outcome; and bias in selecting the reported result. Study quality was independently rated at the individual study level by two authors (LJTS and AS or LJTS and RA) using the Cochrane levels of evidence ratings and RoB2. Tables were organized by the first author’s last name, and extracted data were synthesized.

## Results

The search process results of our systematic review are presented in Figure 1. This review focused on a total of 9 trials^14-22^ of the effects of traditional East Asian medicine (TEAM) on menopausal transition symptoms (hot flashes, sleep, mood, pain, cognitive concerns), representing a total of 811 participants from 7 countries. One trial examined two interventions separately^18^. The interventions tested included acupuncture, acupressure, Chinese herbal medicine, and moxibustion. Moxibustion is a therapeutic application of heat that involves burning mugwort *(Artemesia Vulgaris, Ai Ye*) and holding it above the skin, or directly on the skin, depending on the technique used. Results of efficacy are listed in Table 1. The data table, including statistically significant differences and intervention details, is available in Appendix 1.

**Figure 1.**
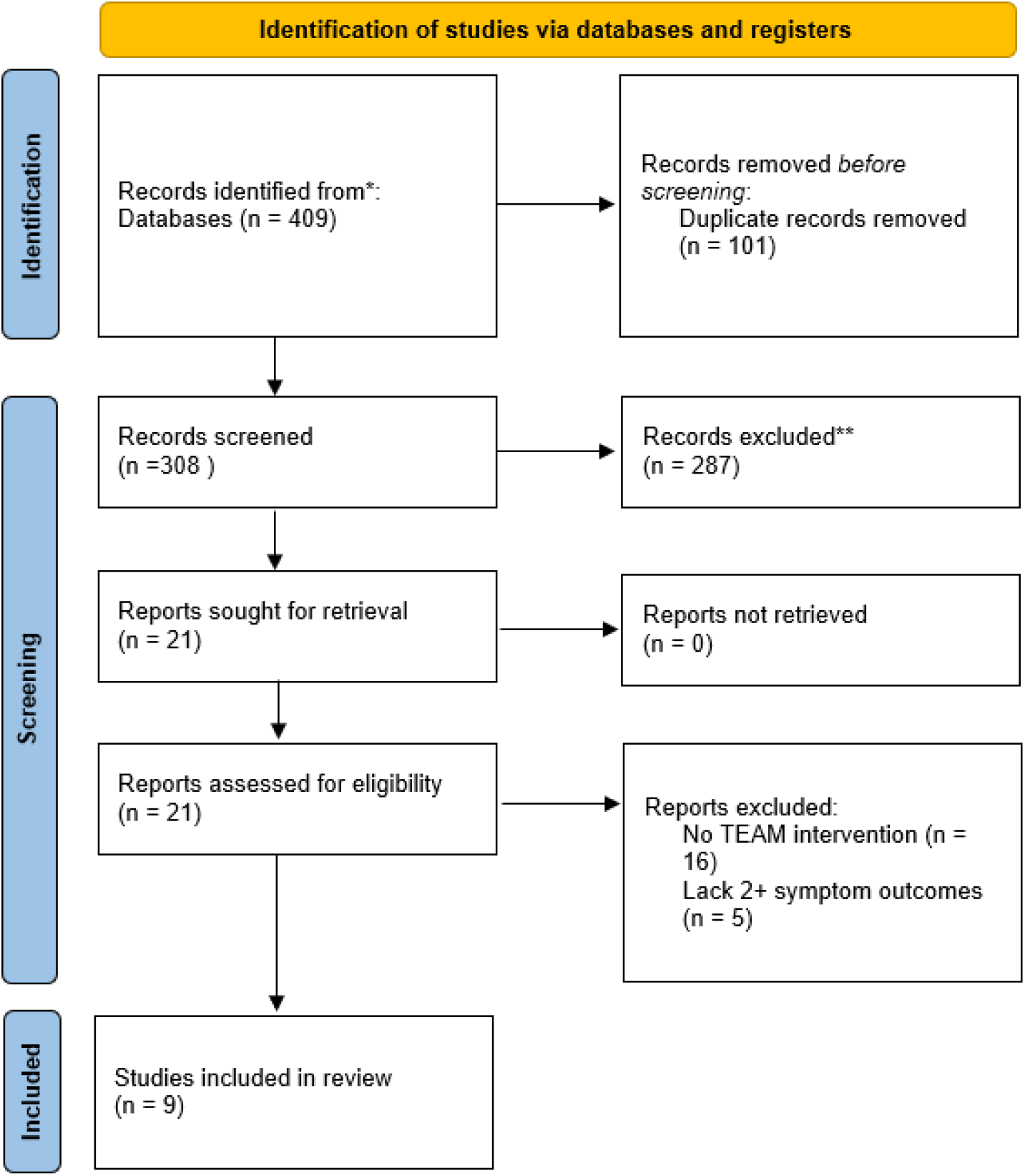
Prisma 2020 flow diagram

**Table 1.**
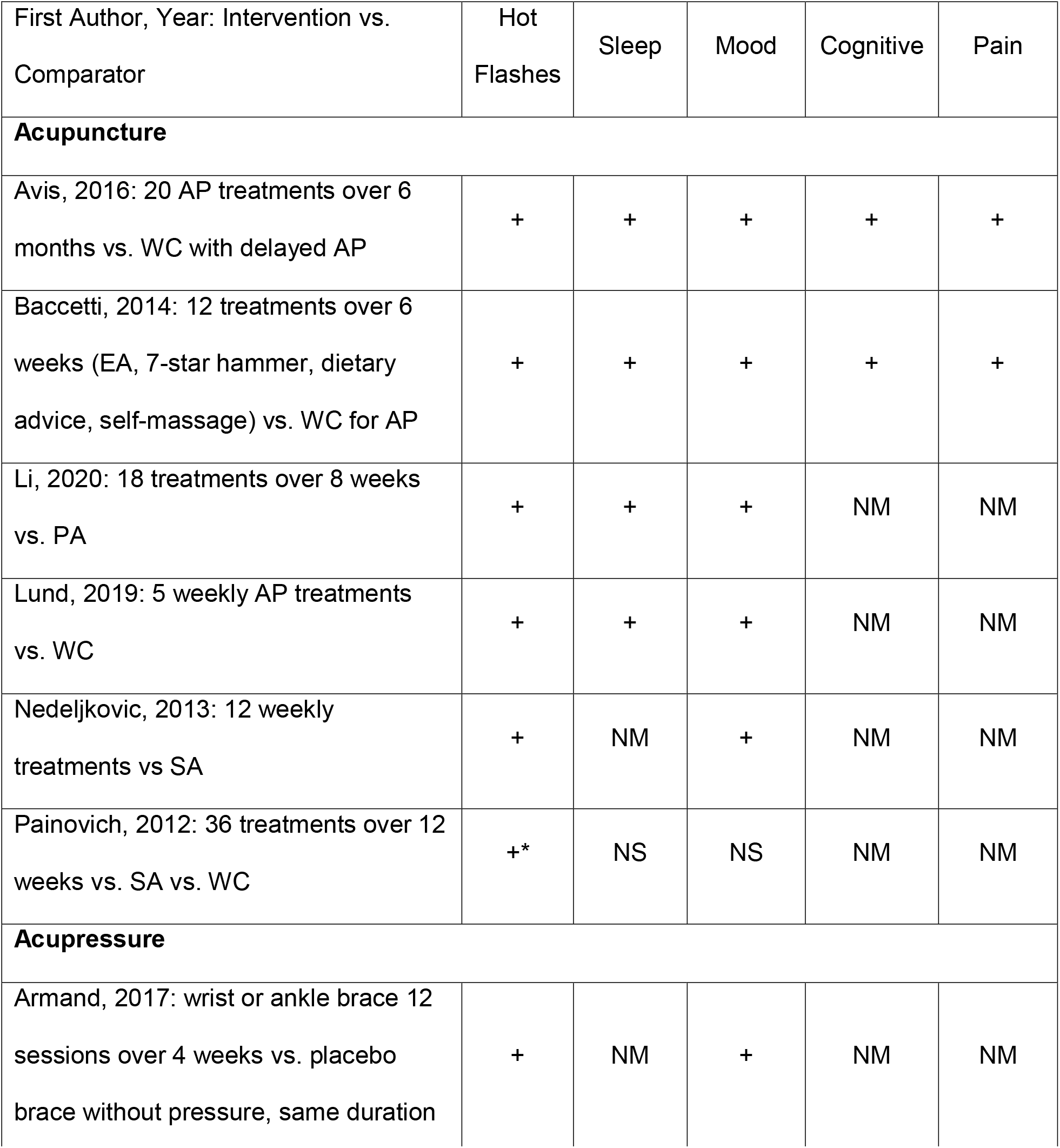
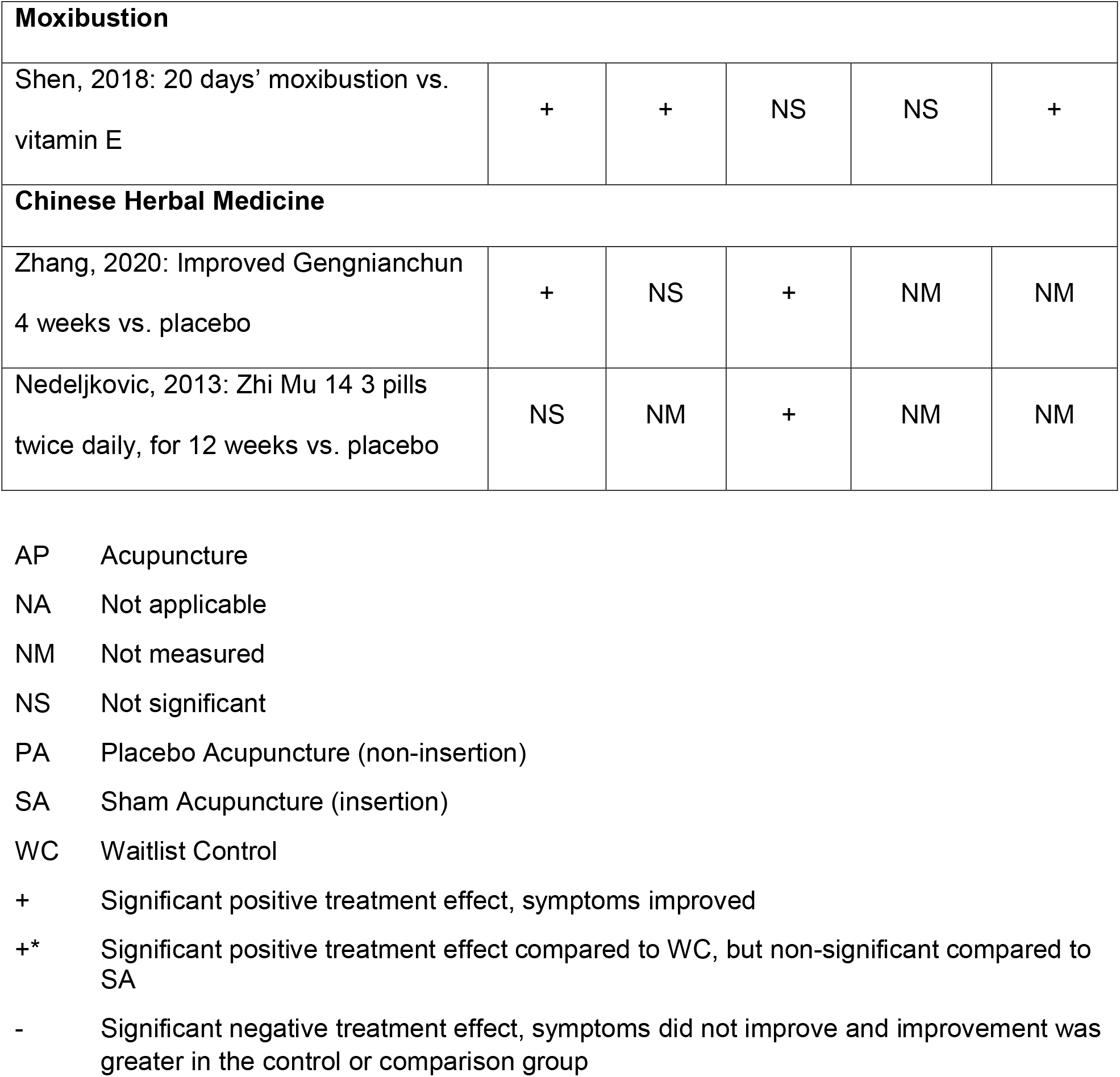
Measure of Efficacy

### Acupuncture, Hot Flashes and Co-Occurring Symptoms

As seen in Table 1, acupuncture treatment was effective in significantly decreasing hot flash symptoms in the six trials reviewed^1-6^. Acupuncture treatment was delivered in 5-36 sessions over 5-12 weeks. Each study used varying acupoints; four of the studies used all or some acupoints chosen according to the individual participants’ TEAM differential diagnosis. Acupuncture treatment resulted in an improvement in pain, sleep, mood, and cognitive symptoms when measured^1-5^. Painovich and colleagues’ study was the single study that reported non-significant improvement in sleep and mood symptoms when measured.^6^ Four of 5 studies that measured sleep reported statistically significant improvement, and 5 of 6 studies reported statistically significant improvement in mood. The effects of acupuncture were maintained at six months post-treatment in the study by Avis and colleagues^1^.

### Acupressure, Hot Flashes and Co-Occurring Symptoms

A significant decrease in hot flashes was observed in the single study analyzed, as seen in Table 1 ^7^. Acupressure treatment was delivered via wearing an acupressure bracelet three times per week for four weeks to analyze the effects of acupressure on 3 acupoints (PC 6 - *shenmen*, SP 6 - *sanyinjiao*, and KI 7 - *fuliu*). Significant improvement in mood was also reported. Changes in sleep, pain, and cognitive concerns were not measured.

### Moxibustion, Hot Flashes, and Co-Occurring Symptoms

Hot flashes, sleep problems, and pain were significantly decreased by moxibustion treatment in the single study analyzed, as seen in Table 1^8^. Four 5-day courses of moxibustion were performed bilaterally at Kidney 3 (*shenshu*) for 15-minute sessions.^8^

### Chinese Herbal Medicine, Hot Flashes and Co-Occurring Symptoms

Chinese herbal medicine (CHM) significantly improved hot flash symptoms in 1 of the 2 studies assessed, as seen in Table 1. The study by Zhang and colleagues noted the use of an I-GNC formula, while the study by Nedeljkovic and colleagues used a standardized CHM preparation of Zhi Mu^5,9^. Chinese herbal medicine had a non-significant effect on sleep symptoms when measured. Both studies showed a positive change in mood symptoms. Pain and cognitive effects were not analyzed by either study, and sleep was measured in 1 of the 2 studies, with non-significant results.

Overall, TEAM was beneficial for *hot flashes and sleep* in 4 acupuncture studies and 1 moxibustion study. TEAM was beneficial for *hot flashes and mood* in 5 acupuncture studies, 1 acupressure study, and 2 CHM studies. TEAM was beneficial for *hot flashes and cognitive concerns* in 2 acupuncture studies. The effects of TEAM on *hot flashes and pain* were beneficial in 2 acupuncture studies and 1 moxibustion study. The studies by Painovich and Nedeljkovic (CHM) had a significant effect on a single symptom, hot flashes and mood, respectively.

### Risk of Bias and Study Quality

Cochrane levels of evidence ratings were noted, and the risk of bias in RCTs was assessed using the Revised Cochrane risk-of-bias tool for randomized trials (RoB2).^13^ Both are described in Table 2. All nine trials are Level I evidence. Of the 6 acupuncture studies, 4 were low risk, 1 was of some concern, and 1 was high risk. The acupressure study was of some concern and the moxibustion study was appraised as high risk of bias. One of the two Chinese herbal medicine studies was of some concern and one was deemed low risk regarding bias. Please also refer to the RoB2 Plots (Figure 2).

**Figure 2.**
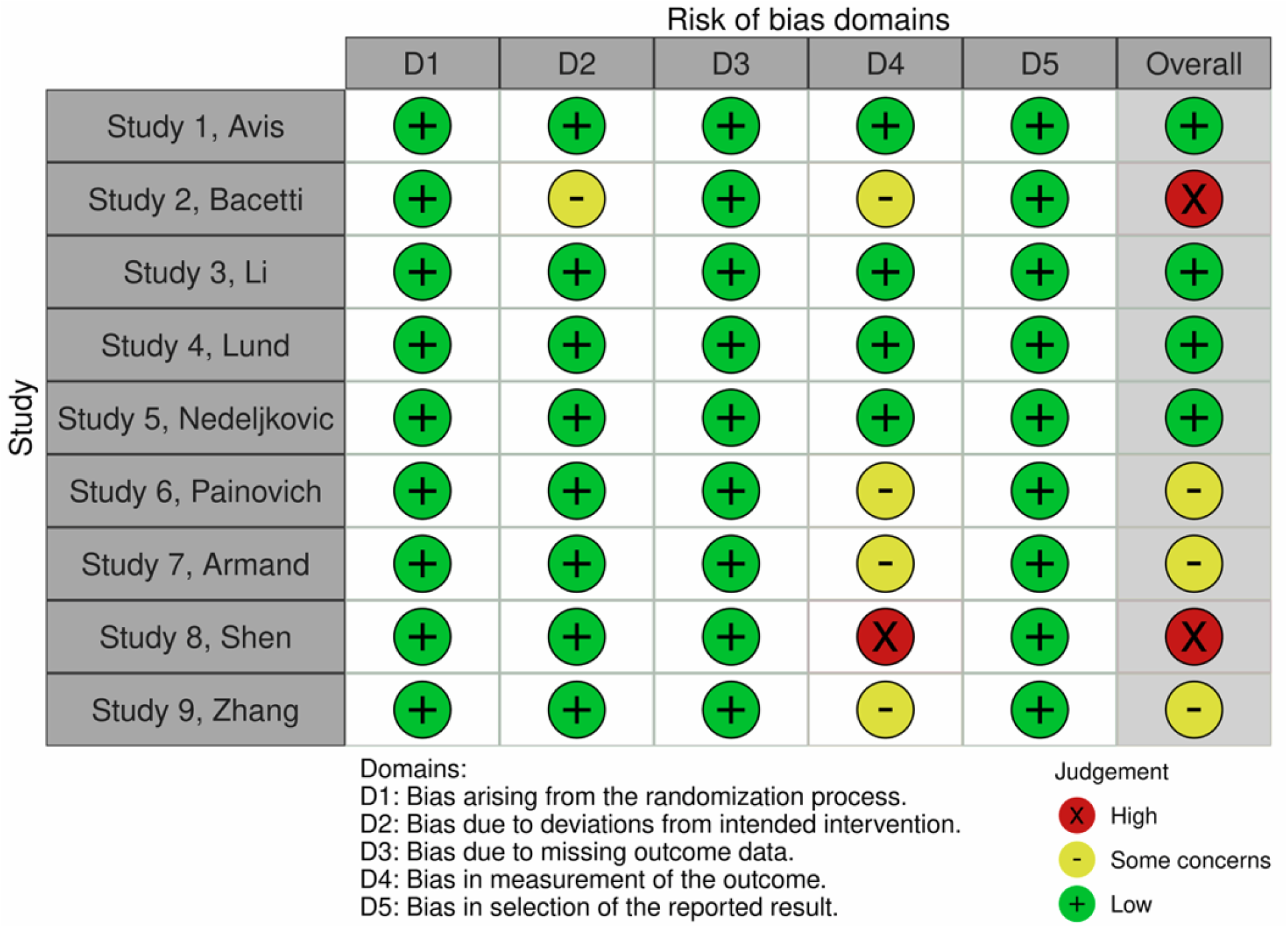
Risk of bias assessment

**Table 2.**
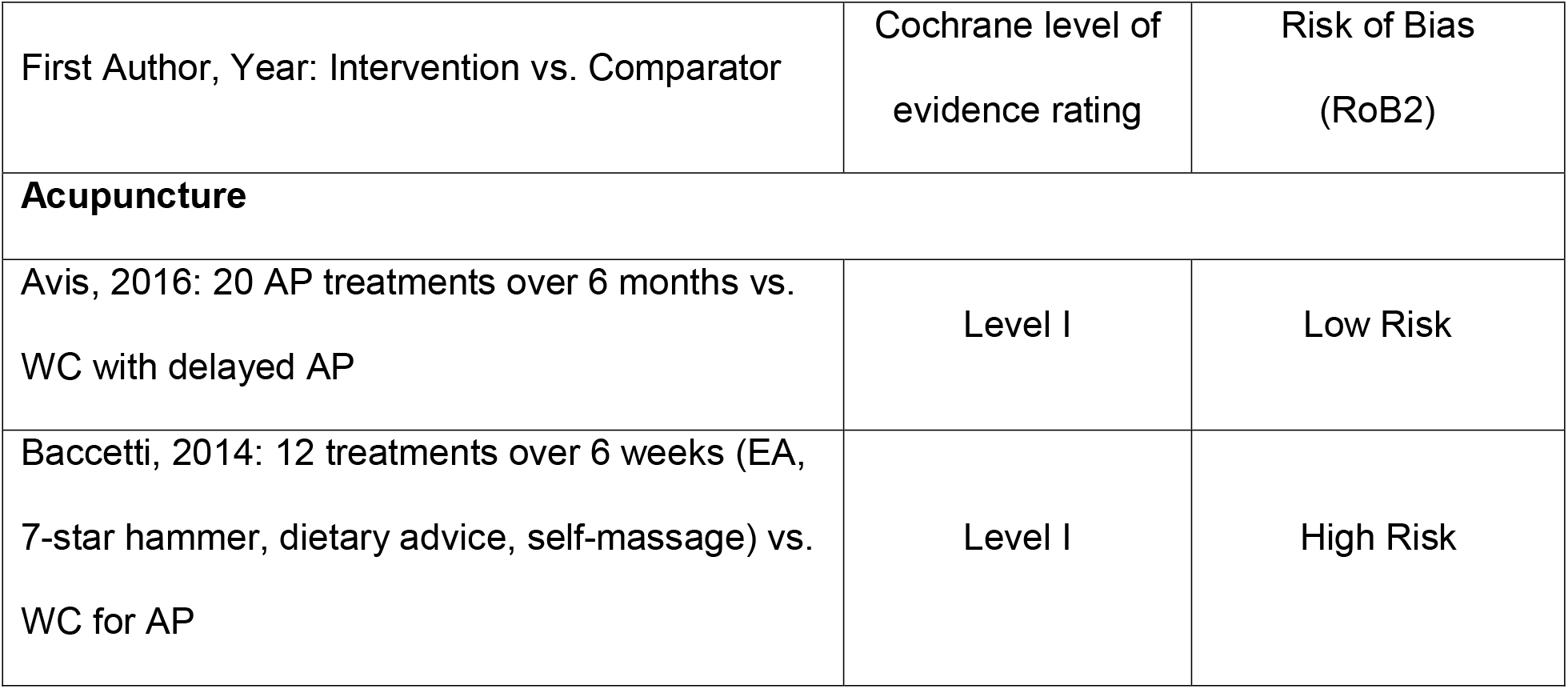
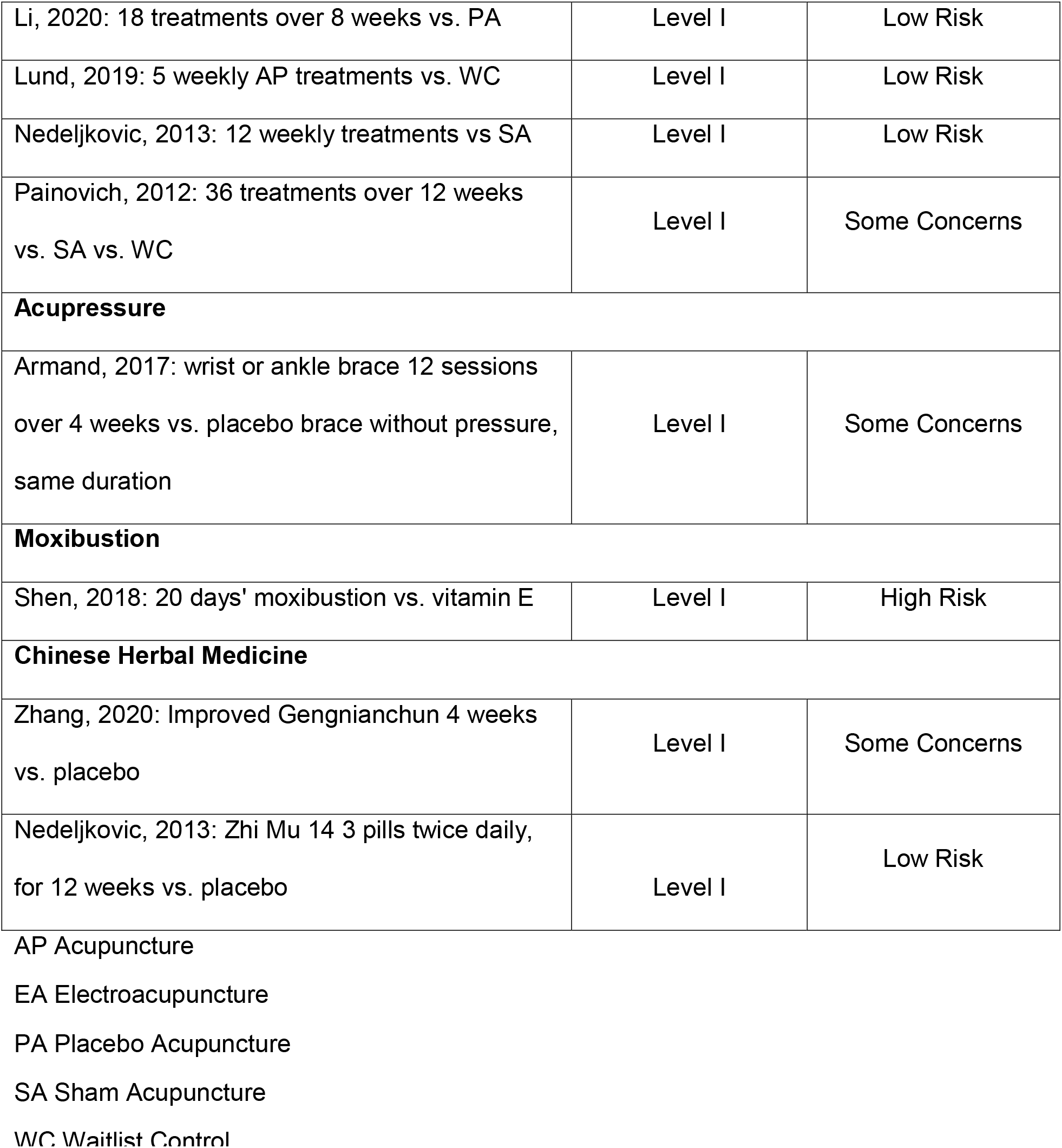
Risk of Bias and Study Quality

### Adverse Events

Adverse events (AEs) were appraised for each trial and are summarized in Table 3. With respect to acupuncture trials, three studies did not report out on AEs^14,15,19^. Three trials reported AEs that were mild (bleeding, pain at needle insertion location), and no serious AEs were reported. The study of acupressure did not report out on AEs^20^. The moxibustion study reported no serious AEs and a minor AE, thirst, occurred for one participant. Among the two studies of Chinese herbal medicine, one study reported no serious AEs or AEs^22^, and one study reported one serious AE, dysplasia of squamous epithelium, which was thought not to be related to the study trial medication^18^.

**Table 3.** Trials of traditional East Asian medicine for hot flashes and associated symptoms: Adverse events and serious adverse events

| Name, year | Adverse events |
| --- | --- |
| **Acupuncture** |  |
| Avis, 2016 | NR |
| Baccetti, 2014 | NR |
| Li, 2020 | Acupuncture AE: (1 bleeding and 1 pain); Sham group AE (1 pain).<br>All AEs were mild and all participants finished the trial. |
| Lund, 2019 | Mild potential AEs were reported by 4 participants; no severe AEs |
| Nedeljkovic, 2013 | Sham group reported a mild AE: pain sensation at needle insertion |
| Painovich, 2012 | NR |
| **Acupressure** |  |
| Armand, 2017 | NR |
| **Moxibustion** |  |
| Shen, 2018 | No serious AEs. Minor AE: 1 person felt thirsty each time after<br>moxibustion and was advised to drink more water. |
| **Chinese Herbal Medicine** |  |
| Zhang, 2020 | No AEs or Serious AEs in either group. |
| Nedeljkovic, 2013 | Serious AE: 1 participant in the verum CHM group, dysplasia of<br>squamous epithelium at the final gynecologic examination. The serious<br>AE is not thought to be related to the trial medication. |

Notes:
AE: Adverse Event
AP: Acupuncture
CHM: Chinese herbal medicine
NR: Not reported
SA: Sham Acupuncture (penetrating)
TCM: Traditional Chinese Medicine

## Discussion

Based on this review, acupuncture, acupressure, moxibustion, and Chinese herbal medicine all show promise in treating hot flashes and sleep problems and hot flashes and mood problems. Acupuncture is a promising treatment of hot flashes and cognitive concerns and hot flashes and pain. Moxibustion appears to provide promising treatment of hot flashes and pain. Regarding the safety profile of these interventions, four studies reported mild adverse events such as thirst, bruising, tenderness at the acupuncture needle insertion site. One study of the nine reviewed reported a severe adverse event that was thought to be unrelated to the study. However, four manuscripts did not report on adverse events. The reported adverse events are summarized in Table 3. Bias was appraised, and studies were primarily with low or some concerns of risk of bias. Therefore, these findings can be appraised for clinical practice, with the safety profile being of utmost concern to clinicians and patients alike. Overall, these interventions demonstrate safety, with low rates of adverse events and only one serious adverse event thought to be unrelated to the study intervention. These findings align with a prospective study including 229,230 patients who received an average of 10 acupuncture treatments, of which 8.5% reported minor adverse events such as bleeding or hematoma^23^. In terms of efficacy for individual symptoms, hot flash improvement was statistically significant in 9 of 10 studies; sleep was statistically significantly improved in 5 of 7 studies measuring sleep outcomes. Mood improved significantly in 8 of 10 studies, while cognitive concerns improved in 2 of 3 studies. Finally, pain was significantly improved in 3 of 3 studies observing pain outcomes. Referring patients to a licensed acupuncturist is prudent, with a favorable safety and efficacy profile. A recent umbrella systematic review and meta-analysis also supports the use of acupuncture for reducing vasomotor symptoms^24^.

Four reviews of TEAM for MT symptoms have been published in the last several years. While each review examined a single intervention for a single primary outcome, the findings of the present review are in alignment with findings of these reviews. He et al.^25^ reviewed 15 RCTs trialing acupuncture against hormone therapy (HT). Compared to HT, acupuncture was significantly associated with a decreased clinical effective rate and decreased Kupperman Index scores. Acupuncture demonstrated very few adverse events; additionally, acupuncture was associated with significantly decreased breast tenderness and gastrointestinal reactions compared to HT. Qin et al.^26^ summarized the evidence and safety of acupuncture and also shared their clinical experiences treatingg outpatient MP women in Beijing. They concluded that acupuncture improves Kupperman Index scores, hot flashes, and pain with few side effects and posit that acupuncture improves symptoms via neuroendocrine-immune systems. Ebrahimi et al.^27^ reviewed 145 articles on herbal interventions for menopausal symptoms and concluded that various individual herbs, including licorice, valerian, soy, sage, and ginseng, showed promise in treating menopausal symptoms. Wang et al.^28^ reviewed 16 trials of Er Xian Tang which is a Chinese herbal formula consisting of six herbs for menopausal symptoms. Er Xian Tang plus HT reduced Kupperman index scores compared to HT alone, but they found the herbal formula versus HT was non-significant. Er Xian Tang significantly reduced total hot flash scores, total MRS scores, total MENQOL scores compared to placebo.

There are several limitations to this review, including the small number of trails that met our criteria for inclusion. Second, the trials are small to moderate, with sample sizes ranging from n=33 to n=209 (mean=56). An additional limitation is the heterogeneity of measures used, so it was difficult to compare findings across studies.

Recommendations for future research include the conduct of pragmatic trials of real-world traditional East Asian medicine, which as is currently practiced typically involves concurrent use of acupuncture, cupping, tui na, Chinese herbal medicine and moxibustion. The selection of interventions is made by clinician decision according to each individual patient’s presentaiton. Thus, demonstrating clinical effectinvess in a pragmatic study would provide essential data to inform broad implementaiton of these interventions in health systems. To date, the AIM study is one of the few pragmatic studies of acupuncture. Strengths include the randomization of 209 participants to receive up to 20 treatments in a 6-month duration, or a wait-list control to receive up to 20 treatments in the second 6-month period. However, while the study allowed concurrent use of moxibustion or electrical stimulation, other modalities such as Chinese herbal medicine, tui na, cupping, or the provision of lifestyle advice and self-care such as self-acupressure were not allowed. Thus, a whole systems TEAM trial designed to measure changes in symptom clusters would add essential literature on the multi-component intervention that is TEAM.

## Conclusions

We found that acupuncture, acupressure, moxibustion, and Chinese herbal medicine are associated with a significant reduction in hot flashes and sleep problems and for hot flashes and mood problems. In addition, each intervention was associated with a low rate of adverse events. Future pragmatic research is needed to study the concurrent use of TEAM interventions’ to treat symptom clusters in the provision of whole person-focused care.

## Data Availability

All data produced in the present work are contained in the manuscript

## Footnotes

1 Sidebar: we write the opening of the introduction section with an inclusive voice indicating that ‘all individuals with a uterus’ will experience the menopausal transition, including transgender and gender-fluid individuals. We subsequently use the term ‘women’ since the vast majority of those with a uterus self-identify as a woman.

