## Appendix 1 for "A systematic review of the effects of traditional East Asian medicine on symptom clusters during the menopausal transition"

**Appendix 1**. Trials of traditional East Asian medicine for hot flashes and co-occurring symptoms: study population, design, interventions, outcomes, and results

|  | **First Author, Year, Study Location** | **Study Population Characteristics, Sample Size** (Screened, Enrolled, Competed, Followed) | **Study Design** | **Main Intervention & Comparison/Control** | **Outcome Measures: Hot Flashes** | **Outcomes Measured: Other Symptoms** | **Results: Hot Flashes** | **Results: Other Symptoms** |
| --- | --- | --- | --- | --- | --- | --- | --- | --- |
| **Acupuncture** (n=6 studies) | | | | | | | | |
| 1 | Avis, 2016 (AIM Study), NC, USA | Include: PeriMP and PostMP, age 45-60. ≥4 VMS per day; surgical MP ok.  Exclude: initiation/change in dose of any treatment for VMS in the last 4 weeks, initiation/change in dose of an anti-depressant in the last 3 months, received acupuncture for any indication in the prior 4 weeks or having received acupuncture from one of the study acupuncturists in the prior 6 months, self-reported health status as poor or fair on the telephone screener, or a diagnosis of hemophilia.  209 enrolled; (170 ACU, 39 control); 33 dropped out; 176 completed; 89% retention at 6 months, 84% retention at 12 months | Pragmatic 2-site RCT with 2 groups (acu and waitlist control).  Block randomization. | Acupuncture Group: Sterile, disposable acupuncture needles were inserted to 0.5-3 cm depth; De Qi sensation attempted; No duration restrictions, or on the application of other modalities (moxibustion, or manual or electrical stimulation of needles); prescribing Chinese herbal remedies was not permitted.  Control Group: No treatment for 6 months then above acupuncture from months 6-12 | HF: frequency, severity Daily Diary of Hot Flashes (DDHF). 4- point scale (mild to very severe). VMS index score (the sum of the number of VMS multiplied by each level of severity). | Sleep, Mood, Cognitive, Pain, and QoL: Women's Health Questionnaire (WHQ) measures physical and emotional health; depressed mood, somatic symptoms, anxiety/fears, vasomotor symptoms, sleep problems, sexual behavior, menstrual symptoms, memory/concentration, and attractiveness  Sleep: Pittsburgh Sleep Quality Index (PSQI) 45 and the PROMIS short form Sleep Disturbance measure  Mood: Depression: Center for Epi-demiologic Studies Depression scale, short form (CESD-10); Anxiety: General Anxiety Disorder (GAD-7), and the PROMIS short form Anxiety; Perceived Stress Perceived Stress Scale (PSS)  QoL: VAS and the Physical and Mental Health Component scores of the Medical Outcomes Study (MOS) 36-Item Short Form Health Survey (SF-36 | HF: significant decline (p<0.001; 36.7%) in VMS frequency among acupuncture group at 6 months; frequency increased by 6% in control group at 6 months; VMS frequency change observed at 3 weeks and significantly different form control (p=0.01) with decreases of 23.7% and 2.4% of baseline frequency in the acupuncture and control groups; reduction from baseline in the acupuncture group at 12 months was 29.5% (p<0.001); control group began receiving treatments at 6 months; between 6 and 12 months, VMS frequency in this group decreased to 31.0% of baseline (p<0.001); maximum reduction in hot flashes occurred at Week 7 (median of 8 acupuncture treatments); Following Week 8, the median cumulative number of treatments continued to increase to a median of 18 at week 26, whereas the percent change in VMS frequency did not. The control group used a median of 16 treatments (months 6 to 12). | WHQ Mood, Cognitive, Pain, and QoL: significantly fewer symptoms in acupuncture compared to control group on the WHQ: vasomotor (p<0.001), Anxiety (p=0.001 @ 2 m), Somatic (p<0.001 @ 2m & 6m), Memory (p=0.001 @ 6m)  Sleep: acupuncture group reported significantly fewer sleep problems on all three measures: the Pittsburgh sleep score (p<0.001 @ 6m); sleep domain of the WHQ (p<0.001 @ 2m & 6m); and PROMIS sleep measure (p=0.002 @ 6m) as compared to women in the control group.  At 12 months, the effects of acupuncture were maintained in the acupuncture group for all of these outcomes.  Control group showed similar improvement on these outcomes and in addition, had reduced depressive symptoms from 6 to 12 months. |
| 2 | Baccetti, 2014, Tuscany, Italy | Women ages 45 to 56 years (mean 51 yrs) experiencing spontaneous MP (ameno ≥ 12 months but ≤ 24 months) with 3+ episodes of HF daily; Exclude: HT within the last month, use of medication for tx of MP, and MP induced by surgery or chemotherapy.    140 women volunteered; 100 first volunteers enrolled.  of 90% Italian ethnicity and all residents of Tuscany; Divided into 2 groups of 50 participants; Sociodemographic factors, alcohol intake, thyroid disease, general health (<0.001), and health in relation to menopause (<0.001) did not significantly differ between groups; 82 followed to completion (41 participants per group at end of tx and 4 months post tx). | RCT with 2 parallel arms. Participants were randomized by envelopes containing group designation.  Group A: TCM diet, self-massage, and electro-acupuncture tx after enrollment  Group B: TCM diet and self-massage at enrollment plus electro-acupuncture tx @ 6 weeks after enrollment | Intervention:  **Acupuncture:** 2 sessions/week for 6 weeks; performed depending on energy diagnosis according to the theory of yin and yang, the Law of Five Phases, and syndromes prevalent in menopause diagnosis. 7-star hammer (plum blossom) in the dorsal region C7–T5 (5 minutes) followed by electro-acupuncture at GV 23 (23 VG shangxing) towards the nose; CV 22 ( 22VC tiantu), BL 2 (2V zanzhu), LI 11 (11GI quchi), LI 4 (4GI hegu); (b) tonification with electrostimulation (40 cycles/ sec) directing the needle toward the groin on SP 10 (10 Rt; Xuehai) and SP6 (6 Rt sayinjiao); and (c) tonification with the needle in the direction of the energetic circulation on GV 20 (20 VG Baihui), CV 4 (4VC guanyuan), CV 6 (6 VC qihai), ST 37 (37 E shangjuxu), and LR 3 (3 F taichong). Acupuncturist added predefined points as needed: deficiency of energy [BL 23] and hot flushes during the night [Ht 3].  **Self-massage:** participants trained in 30-minute self-massage of areas containing acupuncture points (forehead, tempo parietal area, top of head, ear area, lower back area, abdomen, feet, and ankles plant) according to the theory of Tuina Chinese massage  **Diet:** women were classified according to the prevalence of yin and yang type and the Law of Five Phases and prescribed a diet regimen according to finding (Table 1).  Control: control group received the same diet recommendations and self-care; received electro-acupuncture @ 6 weeks after enrollment. | HF and sudden sweating: daily frequency measured; Group A at 2 weeks pre tx, end of tx and 4 months post tx; Group B at 2 weeks pre tx, start of tx, and 4 months post tx | Vaginal dryness, memory loss, genital itching, urinary tract problems, and skin changes: measured by intensity scale rating (1 = no; 2 = slight; 3 = moderate; and 4 = considerable, very marked, serious, intense, or unbearable) measured  Group A at 2 weeks pre tx, end of tx and 4 months post tx; Group B at 2 weeks pre tx, start of tx, and 4 months post tx  Sleep disorders, irritability, bone pain, feeling depressed, headache, chest pain, breast tenderness, and genital bleeding: daily frequency measured  Group A at 2 weeks pre tx, end of tx and 4 months post tx; Group B at 2 weeks pre tx, start of tx, and 4 months post tx | HF and sudden sweating: Group A (1.4 +/- 0.8) showed statistically significant mean improvement in frequency compared with women in group B (0.1 +/- 0.6) for hot flushes (p<0.001) at the end of tx. Results at 4 months post tx did not differ significantly for group A (p=1.00) or B (data not given). For group B statistically significant improvement was recorded between start of tx w/o acupuncture and start of tx with acupuncture. | Group A showed statistically significant mean improvement in frequency and intensity score compared with women in group B for sudden sweating (<0.001), sleep disorders (<0.001), irritability (<0.001), bone pain. (<0.001), feeling depressed (<0.001), headache (<0.001), chest pain (<0.001), memory loss (<0.001), vaginal dryness (<0.001), skin changes (<0.001), urinary tract problems (<0.001), and genital itching (<0.001) at the end of tx. At 4 months post tx group A showed worsened symptoms of vaginal dryness, skin changes, and vaginal itching; only vaginal dryness was statistically significant (<0.001). |
| 3 | Li, 2020, Shanghai, China | 84 perimenopausal women, aged 45-60 (mean 52.5 years), experiencing insomnia were screened. 81 completed the study, 106 screened, 22 excluded; 5 declined to participate and 17 were not postmenopausal or in the menopausal transition, or were using HT drugs. 2 women in acupuncture group & 1 in sham group dropped out for personal reasons.  Include: Diagnosis met ICSD-3 (International Classification of Sleep Disorders) criteria, Age 45 to 60 years old, Amenorrhea for at least 6 months, Insomnia lasting at least 3 months, Total score > 5 on Pittsburgh Sleep Quality Index (PSQI), TCM differen- tiation types (kidney Yin deficiency and kidney Yang defi- ciency), Willingness to participate and be randomly assigned to 1 of the groups, Signed informed consent.  Exclude: Ongoing hormone replacement therapy and/or antidepressant drugs, Induced amenorrhea due to surgery, Any serious physical disease, Insomnia caused by other external factors, PSQI>15, Use of hypnotic medication, except Estazolam, Received acupuncture tx for climacteric syndrome in the last 6 months, Participated in any other clinical trial in the last 6 months, Not capable of understanding the trial or providing responses for the outcome measurements. | Semi-standardized, block randomized sham-controlled trial with 2-parallel-group, blinded (except acupuncturists). Random allocation lists were generated by an independent statistician (WZ) using SAS 9.4. The participants who met the criteria were randomly assigned to either acupuncture or sham acupuncture. The random allocation details were concealed in opaque envelopes.  Only acupuncturists knew the tx allocation. Participants and other relevant researchers (the principal investigator (PI), data analysts, outcome assessors, and statisticians) were kept blinded to the group allocation.  Participants already receiving Estazolam (1–2mg) tx continued use for the intervention period. Participants were told that they had equal odds of receiving real or “placebo” treatment and were free to withdraw from the study at any time. Acupuncturists (SSL and ZQW) were trained professionally for administering this treatment. | 18 sessions of real or non-invasive placebo (sham) tx over the course of 8 weeks (3 times per week for 4 weeks, twice per week for 2 weeks, once per week for 2 weeks). --Patients were placed in separate quiet spaces lying in a supine position and received 30-minute tx based on the diagnosed TCM.  Acupuncture: tx consisting of 10 acupuncture points, 8 main fixed points plus 2 additional points selected by researchers based on the patient’s syndrome differentiation. The main points include Baihui (GV20), shenting (GV24), yintang (GV29), qihai (CV6), guanyuan (CV4) and bilateral anmian (EX-HN22), sanyinjiao (SP6), and shenmen (HT7). Additional points include mingmen (GV4) and shenshu (BL23) for kidney yang deficiency and taiXi (KI3) and fuliu (KI7) for kidney yin deficiency. Sterile and disposable (0.25 × 40 mm and 0.30 × 40 mm in length; Jia Jian, China) needles were inserted into the skin to the depth of 10–30 mm and manipulated manually (manipulation technique included lifting, thrusting, and rotating) until the patient reported needling sensations (Deqi sensation). GV20 and GV29 were connected to a G6805–2 Multi-Purpose Health Device (Huayi Company), using continuous wave type, frequency at 2.5 HZ, and intensity of 4–5 mA. Needles were retained for 30 min before removal. Conversation between acupuncturists and patients was minimal to avoid nonspecific tx effects.  Sham acupuncture: Participants received Streitberger Placebo needle, a non-invasive placebo device. The needle shortens into itself once it touches the skin giving the visual impression of insertion into the skin. AG6805–2 Multi-Purpose Health Device was connected to the GV20 and GV29, w/o electrical pulse. Needles were retained for 30 min before removal. | Climacteric symptoms and quality of life: Vasomotor, psychosocial, physical, and sexual assessments measured by the Menopause Quality of Life (MenQoL) scales at week 0, 4, week 8 and at the post-tx follow-up visits at week 12 and week 20  HF NOT MEASURED ALONE | Sleep: Change in the Pittsburgh Sleep Quality Index (PSQI) btw baseline & end of tx (week 8) results.   **PSQI is a self- rating questionnaire resulting in a global score between 0 and 21, which consists of 7 sub-scores (sleep quality, sleep onset latency, sleep duration, sleep efficiency, sleep disturbances, daytime dysfunction and use of sleep medication).  ***PSQI global score >5 indicates a poor quality of sleep. PSQI were assessed before random assignment (baseline), during tx (week 4), at end of tx (week 8; primary time point), and during follow-up visits (week 12 and week 20).  Sleep Parameters: recorded in the Actigraphy (included TST: total sleep time; SE: sleep efficacy; SA: sleep awakenings; AA: average awakening; WASO: wake after sleep onset) at week 8; sleep disturbance measured by Insomnia Severity Index (ISI) at week 8.  Anxiety + depression: measured by Self-Rating Anxiety Scale (SAS) and Self-Rating Depression Scale (SDS) btw baseline and week 8 only.   At the end of tx, participants were asked to assess which tx they received (acupuncture group, simulated acupuncture treatment group, unsure). | MenQoL dimensions (Table 2):  Vasomotor: Week 4: No statistical significance between the 2 groups (P =0.066). Other weeks showed statistical difference between the 2 groups Week 8: P <0.001 Week 12: P <0.001  Week 20: P <0.001 Physical: All weeks showed statistical significance. Week 4: P= 0.021  Week 8: P <0.001  Week 12: P <0.001  Week 20: P <0.001  Psychosocial and sexual: No statistical difference between groups. | PSQI Score:  Baseline: Statistical significance difference in mean change in PSQI between acupuncture and sham acupuncture group (P <0.001); Table 2; Figure 2). Week 4: No statistically significant difference between acupuncture group and sham acupuncture group (P =0.084). Week 12 and 20: statistically significant differences in the PSQI score between the 2 groups (P <0.001) and (P <0.001), respectively.Acupuncture group reported lower post tx PSQI scores than the sham acupuncture group. The acupuncture group showed a greater reduction in PSQI score compared with the sham acupuncture group (P <0.001).  Table 3 shows the 7 components of PSQI. The aspects of Sleep Duration, Sleep Quality, Sleep Disturbance and Habitual Sleep Efficiency showed the significant difference between 2 groups.  Sleep actigraphy: After tx, there were a significantly higher TST (P =0.007) and SE (P =0.023) in acupuncture than sham.  AA was significantly lower in acupuncture (P =0.011). No significant differences in SA and WASO btw the 2 groups (P > 0.05)  Anxiety + depression: ISI + SAS scores were significantly lower in acupuncture group than sham group (P <0.001 and P =0.007 respectively). |
| 4 | Lund, 2019, Denmark | 70 women experiencing moderate to severe HF aged 40-65 yrs with no significant differences in baseline characteristics. 70 women enrolled, n=34 control and n=36 intervention at week 0, n= 32 control and n=35 intervention at week 3, n=31 control and n=35 (66) intervention at week 6.  Include: 40-65yrs, moderate to severe HF (≥4 on MenQ), intact cognitive function, email address  Exclude: hysterectomy/oophorectomy, alcohol consumption of >21 drinks/week, Rx sleeping pills/sedatives, Hx of breast, ovarian, endometrial, or cervical cancer, Hx of other severe cancer in past 5 years, heart valve disease, insulin dependent/ uncontrolled diabetes mellitus, thyroid disease, under investigation for serious disease, acup tx in past 6 months, pregnant/breastfeeding in past 2yrs, participated in another trial in the 2 weeks before screening, used any of the following in the past 4 weeks: systemic HT, hormonal intrauterine device, antidepressants/antiepileptics, other tx for HF besides inhaled steroids. | Randomized control trial. Control/treatment group allocation generated at 1:1 ratio by computer. Assessors and statistician blinded. | Acupuncture: Participants received 1 acupuncture tx per week for 5 weeks. Standardized Western Medical Acupuncture (WMA) with predefined points (CV-3, CV-4, LR-8, SP-6 and SP-9; LR-8, SP-6 and SP-9 were given bilaterally) performed. Disposable sterile (Plandent) needles, size 0.30×30 mm, were inserted perpendicularly and rotated manually for a few seconds to elicit ‘de-qi’ (needle sensation, a feeling of heaviness around the acupuncture point) and retained for 10 minutes. After each tx, acupuncturist completed a documentation with the date, insertion of each of the needles and whether ‘de-qi’ was achieved.  Control: participants offered one acupuncture treatment after week 5  Participants received MSQ electronically at weeks 0, 3, 6, 8, 11 and 26, follow up at week 3 (intermediate assessment) and week 6 (final assessment). | HF: measured by MSQ mean scores from baseline to week 6 | Differences measured by differences in mean MSQ score from baseline to week 6: -Day & night sweats and general sweating  -Meno. specific sleep problems -Emotional sx -Memory changes -Physical sx -Urinary & vaginal sx -Abd. Sx -Skin & hair sx -Sexual sx -Tiredness  All analyses were based on intention-to-treat analysis. | HF: intervention group MSQ mean scores significantly reduced at  3 weeks (p=0.0002) and 6 weeks (p<0.0001). | In the intervention group, 80% of participants reported a general beneficial treatment effect after 6 weeks.   Statistically significant differences were identified at 6 weeks in the following Secondary outcomes:  Day and night sweating p=0.0056  General sweating p=0.0086  Meno. specific sleep problems p<0.0001 Emotional symptoms  p=0.0008, Physical sx p=0.010  Skin and hair sx p=0.0021  Significant decrease in emotional sx (p=0.0015, skin and hair sx (p= 0.0036) in week 3 |
| 5 | Nedeljkovic, 2013, Bern, Switzerland  INCLUDES AP AND CHM | 63 individuals were assessed for eligibility, 40 participants met the inclusion criteria. An independent data manager carried out randomization by using a computer-generated random allocation sequence. Participants were assigned to TCM AP, sham AP, verum CHM, or placebo CHM group equally (1:1:1:1). 10 TCM AP, 10 Sham AP, 10 Verum CHM, and 9 Placebo CHM participants completed the study. (39 participants)  Include: hot flushes for at least 1 year, at least 20 hot flushes per week during the run-in period, normal gynecologic status, at least 12 months of self-defined amenorrhea or had undergone hysterectomy, body mass index lower than 30 kg/m2, initial score of at least 20 points on the Menopause Rating Scale (MRS) II, follicle-stimulating hormone serum concentration higher than 30 IU/L, and a signed informed consent form  Exclude: HT and/or treatment with TCM and/or any kind of surgical intervention within 12 weeks of recruitment, abnormal genital bleeding, abnormal liver function, bilateral ovariectomy, pregnancy; chronic and/or acute physical diseases and/or mental disorders, abuse of alcohol and/or any other addictive substances, recent or planned phytoestrogen enriched diet, intake of herbal remedies for treating menopausal symptoms, more than 2 weeks of planned absence during the treatment period, simultaneous participation in any other clinical trial. | Four-arm, prospective, randomized trial. AP treatment was sham controlled and single blinded. CHM treatment was placebo controlled and double blinded.  Participants took part in the study for a total of 26 weeks, including a run-in period with diary collection (2wk), a treatment period (12 wk), and a follow-up period after completion of treatment (12 wk).  All study participants completed **questionnaires** at the end of the run-in period (base-line), 4 weeks after the start of treatment, at the end of treatment, and 12 weeks after treatment completion (follow-up). | Participants in the TCM and sham AP groups were scheduled for 12 weekly treatments. On the first treatment session, women received a TCM diagnosis.  TCM AP Group: Stanard points CV-4, GV-20, GB-20, PC-6, ST-36, SP-6, LI-4, and KI-3 needled. All standard treatment points were needled bilaterally except for CV-4 and GV-20, which are located at the median of the body. In addition to this standardized treatment, 7 to 10 supplementary AP points were needled based on a person’s TCM diagnostic category (ie, B kidney yang with spleen yang deficiency [: BL-20, BL-23, SP-9, and CV-6; B kidney yin with liver yin deficiency [: BL-18, BL-23, HT-6, KI-6, and LR-3) or in accordance with the physician’s clinical judgment. No more than 24 points were needled during any treatment. Needles were inserted through the skin to a depth of 0.2 to 1.5 cm, depending on the target site. With each needle insertion, the acupuncturist attempted to elicit a de Qi sensation around the stimulated AP point. The retention time of the sterile, single-use, stainless-steel AP needles was 30 minutes, without any additional stimulation.  Sham AP Group: P needles were inserted superficially, without attempting to elicit a de Qi sensation, into seven bilateral target sites that did not correspond to established TCM AP points (see Table, Supplemental Digital Content 1, which describes sham AP points in detail).  Participants in the verum and placebo CHM groups were scheduled for clinic visits on weeks 1, 4, 8, and 12 of the treatment period. Women in both groups were instructed to take 3 capsules orally with water twice per day (morning and evening) and to report daily medication intake in their hot flush diary.  Verum CHM Group:  Women received Zhi Mu 14, a standardized CHM preparation comprising 14 plant materials (see Table, Supplemental Digital Content 2, which describes the composition of the CHM formula Zhi Mu 14 in detail). The formulation of Zhi Mu 14 is based on two modified classic formulas VB Gan Mai Da Zao Tang and Qing Hao Bie Jia Tang designed to treat hot flushes, night sweats, insomnia, mood swings, irritability, and emotional instability resulting from kidney yin deficiency. The dose of herbal extract granules (3 g/d) administered is equivalent to 15 g of dry herb.  Placebo CHM Group: women received placebo capsules containing starch (Amylum maydis) and caramel as color tracer. This placebo mixture has no known effects on menopausal symptoms.  Both placebo and CHM capsules were identical in appearance and were prepared by Sheng Foong Pharmaceutical Ltd (Taiwan) on behalf of China Medical Ltd (Aesch, Switzerland). | HF: participants **recorded hot flush severity and frequency per week**, an adaptation of the daily hot flush diary. Hot flush severity score was calculated as the sum of the number of self-reported hot flushes multiplied by severity. The applied hot flush severity rating scale scores were as follows: 1 = mild (heat sensation without sweating and disruption of activity), 2 = moderate (heat sensation accompanied by sweating but with no disruption of activity), and 3 = severe (heat sensation accompanied by sweating and disruption of activity). Participants were asked to record the occurrence and severity of each hot flush during the run-in and treatment periods, and on the 4th, 8th, and 12th weeks of the follow-up period of the trial. | Mood: Menopause related quality of life was assessed using the validated MRS II. This self-report instrument comprises 11 items and assesses the presence and intensity of menopausal symptoms on a 5-pointrating scale ranging from 0 (no symptom) to 4(very severe symptom). A **psychological subscale of MRS II represents the domain specific index of severity of climacteric-related psychological complaints** (ie, 0-4 =none tolittle;5-8=mild;9-16=moderate; 17 =severe). The mean reference value for the MRS II psychological subscale in European populations is 2.9. | HF:  Between-group analyses showed that, in comparison with the sham AP group, participants in the TCM AP group reported a significantly greater decline in hot flush severity (P= 0.013) and hot flush frequency (P= 0.016) from pretreatment to posttreatment, but not from pretreatment to follow-up assessment (P’s90.24; Table 2). | Mood:  psychological MRS II subscale score decreased significantly from pretreatment to post treatment (p<0.017) and to follow-up assessment (p<0.005), |
| 6 | Painovich, 2012, Los Angeles, California, USA | 33 peri- and post-meno women over the age of 40 with meno-related VMS including at least 7 HF per day and at least 1 missed menstrual cycle or spontaneous or medically induced meno. No significant demographic or clinical differences between groups (table 1).  Exclude: concomitant illness with expected survival of less than one-year, current substance abuse, known, suspected or planned pregnancy in next year, other concomitant meno tx, participating in acupuncture tx or formal psychological stress management program within the last year, participating in another tx for VMS, unless willing to stop it 4 weeks in advance of participation, HIV infection, chronic or active hepatitis or other bloodborne illness.  60 women enrolled, 27 dropped out, 33 completed. TA and SA groups had 8 drop out. 11 of WC dropped out due to being in no tx group. | Randomized, single blinded (participant), placebo-controlled trial. Participants equally randomized into TA, SA, and WC through a statistically randomized block design.  TA: Traditional acupuncture 3x/week for 12 weeks  SA: Sham acupuncture 3x/week for 12 weeks  WC: Waiting control for 12 weeks | TA: Points included 11 ‘front’ (DU 20, PC 6, HT 7, LIV 3, LI 4, LI 11, KD 3, SP 6, ST 36, REN 17, REN 6,) placed while supine and 7 ‘back’ (DU 14, UB 15, UB 18, UB 20, UB 23, GB 34 and KD 3) placed while prone. Needles were inserted 0.5 to 1.5 inches, manually stimulated to reach ‘de-qi’, secured with adhesive tape (similar to SA group), and retained for 30 minutes. Required to wear eye coverings during tx.  SA: Points proximate to TA site were selected to enhance blinding without being considered active. Disposable needles and plastic tubes were placed on sham points, manipulated without skin penetration and secured with adhesive tape (Does not include time retained). Required to wear eye coverings during tx.  WC: Received no treatment for 3 months. Offered complimentary TA after trial completion. | HF: measured by VSM frequency (ave # of HF per day for 7 days) and VSM severity (mild, moderate, severe, and very severe) using a 7-day HF diary filled out at wk 0 (entry), wk 5, and wk 12 (exit). | Data collected at wk 0 (entry) and wk 12 (exit)  QOL (vasomotor, psychological, physical, and sexual): MENQOL questionnaire  Sleep quality and disturbance: Pittsburgh Sleep Quality Index (PSI) questionnaire used as self-rating to produce one score between 0 and 21, with higher scores indicating worse sleep quality.  Depression: Beck Depression Inventory II (BDI) used to assess intensity of depression.  Anxiety: State-Trait Anxiety Inventory (STAI) used to differentiate between temporary state and general long-standing anxiety | HF: No significant difference in reduction of VMS frequency and severity between the TA and SA groups (exit-entry). Both groups improved compared to the WC group on VMS frequency (p = 0.24) and severity (p = 0.20) (Figure 2). The majority of the reduction of VMS frequency (≥ 86%) and severity (≥78%) occurred by week five of treatment. The SA severity scores remained static after the initial reduction while the TA scores continued to reduce throughout the 3-month period. | QOL (vasomotor, psychological, physical, and sexual): Significant improvement shown in the vasomotor domain (p = 0.04), and a trend towards improvement in the overall MENQOL (p = 0.07) (TA vs. SA vs. WC). No significant improvement in psychosocial (p = 0.16), physical (p = 0.17), and sexual (p = 0.72) domains (Table 2).  Sleep quality and disturbance: No significant difference between entry-exit frequency (p = 0.15) and severity (p = 0.14) (TA vs. SA vs. WC). TA and SA experienced improvement in both.  Depression: No significant difference between entry and exit frequency (p = 0.09) and severity (p = 0.07) (TA vs. SA vs. WC). TA and SA experienced improvement in both.  Anxiety: No significant difference between entry and exit frequency (p = 0.09) and severity (p = 0.07) (TA vs. SA vs. WC). TA and SA experienced improvement in both.  When TA + SA vs. WC, the relationship became significant for improvement of sleep, depression, and anxiety outcomes (Table 3). |
| **Acupressure** (n=1 study) | | | | | | | | |
| 7 | Armand, 2017, Tehran, Iran | 70 menopaused women 48-51 (mean of 49.83 study and 55.09 placebo) years old with sleep disorders in Yaftaba and Shadabad healthcare centers in 2012. No significant difference in mentioned variables between groups.  79 enrolled, 70 completed (4 study group and 5 placebo group dropped out) Three subjects were left out of the study due to their change in residence, one due to spouse’s death, and five for irregularity in administration of intervention or absenteeism in the scheduled sessions.  Include: early complication of menopause such as hot flashes and night sweats (based on questionnaire and daily record form) and anxiety (based on Spillburger questionnaire), reporting ta least 2 hot flashes per day during the day before the study, normal menopause, 12+ months post menstruation (mean 1.77 yrs study and 1.74 yrs placebo)  Exclude: hormone therapy in the past 6 months, taking tranquilizers, Isoflavin complements, herbals, and vitamin E, history of current systemic or chronic diseases, serious events such as acute and serious disease of a family member or their death, a divorce, unexpected events such as an accident in the past 6-12 months, current acute known disease in hands or legs acting as an obstacle for the administration of the technique, addiction and drug abuse, drinking   and smoking, current application of acupressure, development of medical problems during the study, loss of interest to stay in the study, incorrect use of the wrist braces, any diagnosed mental disorders leading to anxiety, and occurrence of miserable events during the study | Randomized control trial. Randomized through clinical trial random allocation software (coded with numbers 1 or 2).  Intervention administered in 2 sessions of 2 hours every 2 weeks and lasted 1 month. At the end of every 2 weeks, the confounding factors and exclusion criteria were rechecked based on number 2 questionnaire. State and trait anxiety were measured by Spillburger questionnaire, based on the supervisor’s indication and literature review. To administer routine menopausal care sessions including counseling and for educating the patient about the importance of lifestyle related changes, health and menopausal complications, measurement of height, weight, BP, breast exam, and Pap smear test were conducted for each group separately on different days (to make the study blind to the subjects), once at the beginning of the first week and another time at the beginning of the third week. | Acupressure: subjects were given two wrist braces with a pressure button and the researcher educated them about the correct method of their usage. They were trained to wear the wrist braces three times a week (every other day) for 15 min once a day on the special points (Shenmen: On horizontal line of the wrist and in internal side of ulnar bone), (Sanyinjiao: 3 cuns superior to the internal malleolus prominence, one finger behind the inner edge of the tibia bone), and [fuliu: In the anteroexternal face of the leg and 2 cuns upper than Taixi point (between internal ankle and calcaneal tenon )].  Placebo: in addition to routine care, the subject was given two wrist braces with a pressure button and was asked to wear the braces in a counter wise form in such a way that the pressure button was laid outward (no pressure was given to the points).  The researcher observed and supervised the subjects while the subjects wore the wrist braces, and solved their problems in wearing them and gave them appropriate feedback. Also, an educational picture showing the exact location of the abovementioned acupoints was given to each subject at the end of each week; the method of wearing the wrist braces by the subjects was checked through a telephone call. | HF: a daily record of hot flashes and night sweats used to measure frequency and severity at baseline, end of week 1, end of week 2, end of week 3, and end of week 4. hot flash severity was scored as minor (1), moderate (2), and severe (3). The total score of hot flash was calculated using the formula: (1 × number of minor hot flash) + (2 × number of moderate hot flash) + (3 × number of severe hot flash). The severity of night sweats was scored as minor (1), moderate (2), and severe (3) and was calculated using the formula: (1 × number of minor night sweats) + (2 × number of moderate night sweats) + (3 × number of severe night sweats). | Mood: Spillburger Anxiety Questionnaire was used to measure state and trait anxiety at baseline, end of second week, and end of fourth week. Scale included very low anxiety (0-20), low anxiety (21-40), high anxiety (41-60), and very high anxiety (61-80). | HF: results showed a significant reduction in study group frequency of hot flashes from BL to week 4 (4.91+/-1.22 to 1.69+/- 0.99; p<0.001) and severity of hot flashes (10.68+/-4.46 to 3.96+/-3.90; p<0.001) and when compared to placebo at 2nd, 3rd, and 4th wk (p<0.001). Results showed a significant reduction in study group frequency of night sweats from BL to week 4 (2.80+/-1.07 to 1.54+/-0.74; p<0.001) and severity of night sweats (1.91+/-0.74 to 1.46+/-1.46+/-0.56; p<0.001) and when compared to placebo at 3rd and 4th wk (p<0.001). | Mood: results showed a significant reduction in study group state anxiety from BL to week 4 (very low anxiety in 11.40% @ baseline, 31.40% @ end of 2nd week, and 65.70% at end of 4th week; p<0.001) and compared to placebo at the end of week 4 (65.70% very low in study; 14.30% very low in placebo; p<0.001). There was no significant reduction in trait anxiety (p>0.05). |
| **Moxibustion** (n=1 study) | | | | | | | | |
| 8 | Shen, 2018, Nanjing, China | 60 pre- and post-meno women between the ages of 40 and 60 years experiencing repeated discomfort or significant loss of adaptive capacity for more than 3 months but are able to maintain normal activities. No significant clinical differences between groups (Table 1; p>0.05)  Exclude: significant organic or mental disease  60 women enrolled, 28 in moxibustion, 27 in control, 5 dropped out, 55 completed | Randomized, control trial.  Participants blinded to tx type until tx began.  Participants equally randomized into moxibustion or control by stochastic computer program.  Moxibustion: mild moxibustion4 5-day courses with 2 days between courses; 28 tx total  Control Group: vitamin E 1 capsule nightly for 28 days; 28 tx total | Moxibustion: mild moxibustion at BL23 (shenshu) bilaterally with 25-g moxa stick (suzhou moxa co.) for 15 minutes per day so the participant felt comfortably warm; 4 5-day courses with 2 days in between courses; 28 tx total  Control Group: took 1 vitamin E capsule (dose not specified) by mouth nightly after their main meal for 28 days; 28 tx total | HF: frequency of hot flashes added to the questionnaire section addressing physical condition to access physiological characteristics of peri-menopausal women  HF NOT MEASURED ALONE | Participant’s physical condition, livings conditions, emotional status, and energy status observed, recorded, and rated on a scale of 1-5 using a questionnaire. physical condition and living conditions: higher scores indicate more severe symptoms  emotional and energy status: lower scores indicate more severe symptoms.  Pain: physical condition (dizziness, dry throat, sore eyes, mouth ulcers, and appetite)  Sleep: living conditions (sleep conditions, defecation, urination, and work ability)  Mood: emotional status (confidence, whether it is easy to get angry, attitude about life)  Cognitive: energy status (work efficiency, energy outside of work, daily life ability)  Sex Hormones: assessment – blood serum levels of FSH, LH, E2, T, P, and PBL*** | HF: physical condition significantly improved in the moxibustion group compared with the control group (P<0.01; Figure 2).  HF NOT MEASURED ALONE | Pain, Sleep, Mood, Cognitive: physical condition and living conditions significantly improved in the moxibustion group compared with the control group (p<0.01; Figure 2). Physical condition, living conditions, and emotional status were improved in pre-menopausal women (p<0.05; Figure 3). Only physical condition and living conditions improved significantly in post-menopausal women (p<0.01). Independent two-sample t tests indicated that physical condition, living conditions (p<0.05), and emotional status (p<0.01) of post-menopausal women were worse than those of pre-menopausal ones.  Sex Hormones: moxibustion group had significantly higher E2 levels (p<0.01) than the control group after treatment (Figure 4), and the moxibustion group E2 levels increased significantly with treatment in pre-menopausal women (p<0.01, Figure 5). Progesterone, E2 levels (p<0.01; Figures 5, 6), and PBL levels (p<0.05; Figure 5) were significantly higher in pre-menopausal women, whereas LH and FSH levels were significantly higher for postmenopausal women (p<0.01; Figure 5). There were no significant differences in testosterone levels. |
| **Chinese Herbal Medicine** (n=2 studies) | | | | | | | | |
| 9 | Zhang, 2020, Shanghai, China | 275 Indviduals screened and randomized in a 1:1 ratio between I-GNC and placebo groups. 137 randomized to I-GNC, 138 randomized to control. 71 I-GNC and 38 placebo participants remained after exclusion criteria for KMI and dropouts.  Include: ages 41 to 60years, experiencing irregular menstrual cycles indicating perimenopause or cessation of menstruation for at least 3 months within the previous 12 months, experiencing hot flushes and having a Kupperman Index (KMI) score of 15 or higher (moderate to severe menopausal symptoms with hot flushes).  Exclude: cancer, cardio-vascular disease, autoimmune system disease, thrombosis, or thrombophlebitis, fibroids (diameter>3 cm) or a pathological cyst of the ovary, concurrent major depressive disorder with symptoms such as anxiety, insomnia, poor concentration, poor memory or mental retardation, or the use of psychiatric or psychotherapeutic drugs within the last 3 months, use of hormones or similar medications that could affect vasomotor symptoms or Chinese herbal medicines, herbal remedies, over the counter medications, or hormone therapies intended to manage menopausal symptoms within the last 3 months, use of anticoagulation medicines within the last 3 months, or pregnancy.  Dropout criteria: natural loss to follow-up, recommendation that the participant withdraw due to poor compliance, use of less than 80% or more than120% of the dosage, and request by the participant to withdraw. | Randomized placebo-controlled trial.  Participants randomly assigned to the I-GNC group or placebo group at a ratio of 1:1. Treatment assignments were not revealed to the participants until the study was completed.  Participants were required to attend a visit at the Obstetrics and Gynecology Hospital of Fudan University every 4 weeks to complete the KMI, Hamilton depression scale (HAMD), Hamilton anxiety scale (HAMA), and Pittsburgh Sleep Quality Index (PSQI) and they were given a 4-week supply of granulated I-GNC medication or placebo at the visit. Participants recorded daily hot flush frequency (24-hour period). | I-GNC Group: Each dose contained 97 g of crude herbal materials extracted into 11.6 g of granules and packaged in an aluminum foil sachet with a medicinal composite membrane inside. The ingredients of the I-GNC formula are shown in Table 1.  Placebo Group: placebo contained 10% I-GNC granules and other inactive ingredients, and the placebo drug resembled the I-GNC granules in taste, dosage, and wrapping.  Participants were instructed to drink one sachet of granules dissolved in 100 mL of warm water once daily. A 4-week supply was dispensed at each treatment visit. | HF: The **frequency of hot flushes** was recorded daily (24-hour period) and was assessed every 4 weeks up to 12 weeks, the endpoint of this trial. | Mood: The **HAMD** scores were classified into four levels of severity (0 to 6) no depression, (7 to 17) possible depression, (18 to 28) moderate or mild depression, and (29 or more) severe depression. The **HAMA** scores were classified into five levels of severity (0 to 6) no anxiety, (7 to 14) possible anxiety, (15 to 21) highly likely anxiety, (22 to 29) obvious anxiety, and (more than 30 points) severe anxiety. Scores recorded every 4 weeks for 12 weeks.  Sleep: PSQI measured every 4 weeks for 12 weeks to evaluate sleep disturbance over the past month, and the total score ranges from 0 to 21. The higher the score, the worse the sleep quality | HF: The mean (SD) frequency of hot flushes per day decreased from 7 (4.55) to 1.2 (1.68) (P<0.01) in the I-GNC group and from 6.74 (3.43) to 3.66 (2.64) (P<0.01) in the placebo group during the 12-week treatment period (P<0.01). Evaluations at 4 and 8 weeks during our trial also showed **significant differences between the two groups (P<0.01)** (Table 3). | Mood: Fisher exact test was used to compare the HAMD, and HAMA scores between the I-GNC group and the placebo group. The changes in the **HAMD and HAMA scores in the I-GNC group were significantly different from those in the placebo group (P<0.05)** (Table 4).  Sleep: There was **no significant difference in the PSQI score** (P= 0.204) at 12 weeks between the two groups. No other significant differences at 0,4, or 8 weeks were observed (Table5). |
| 10 | Nedeljkovic, 2013, Bern, Switzerland  INCLUDES AP AND CHM | 63 individuals were assessed for eligibility, 40 participants met the inclusion criteria. An independent data manager carried out randomization by using a computer-generated random allocation sequence. Participants were assigned to TCM AP, sham AP, verum CHM, or placebo CHM group equally (1:1:1:1). 10 TCM AP, 10 Sham AP, 10 Verum CHM, and 9 Placebo CHM participants completed the study.  Include: hot flushes for at least 1 year, at least 20 hot flushes per week during the run-in period, normal gynecologic status, at least 12 months of self-defined amenorrhea or had undergone hysterectomy, body mass index lower than 30 kg/m2, initial score of at least 20 points on the Menopause Rating Scale (MRS) II, follicle-stimulating hormone serum concentration higher than 30 IU/L, and a signed informed consent form  Exclude: HT and/or treatment with TCM and/or any kind of surgical intervention within 12 weeks of recruitment, abnormal genital bleeding, abnormal liver function, bilateral ovariectomy, pregnancy; chronic and/or acute physical diseases and/or mental disorders, abuse of alcohol and/or any other addictive substances, recent or planned phytoestrogen enriched diet, intake of herbal remedies for treating menopausal symptoms, more than 2 weeks of planned absence during the treatment period, simultaneous participation in any other clinical trial | Four-arm, prospective, randomized trial. AP treatment was sham controlled and single blinded. CHM treatment was placebo controlled and double blinded.  Participants took part in the study for a total of 26 weeks, including a run-in period with diary collection (2wk), a treatment period (12 wk), and a follow-up period after completion of treatment (12 wk).  All study participants completed **questionnaires** at the end of the run-in period (base-line), 4 weeks after the start of treatment, at the end of treatment, and 12 weeks after treatment completion (follow-up). | Participants in the TCM and sham AP groups were scheduled for 12 weekly treatments. On the first treatment session, women received a TCM diagnosis.  TCM AP Group: Stanard points CV-4, GV-20, GB-20, PC-6, ST-36, SP-6, LI-4, and KI-3 needled. All standard treatment points were needled bilaterally except for CV-4 and GV-20, which are located at the median of the body. In addition to this standardized treatment, 7 to 10 supplementary AP points were needled based on a person’s TCM diagnostic category (ie, B kidney yang with spleen yang deficiency [: BL-20, BL-23, SP-9, and CV-6; B kidney yin with liver yin deficiency [: BL-18, BL-23, HT-6, KI-6, and LR-3) or in accordance with the physician’s clinical judgment. No more than 24 points were needled during any treatment. Needles were inserted through the skin to a depth of 0.2 to 1.5 cm, depending on the target site. With each needle insertion, the acupuncturist attempted to elicit a de Qi sensation around the stimulated AP point. The retention time of the sterile, single-use, stainless-steel AP needles was 30 minutes, without any additional stimulation.  Sham AP Group: P needles were inserted superficially, without attempting to elicit a de Qi sensation, into seven bilateral target sites that did not correspond to established TCM AP points (see Table, Supplemental Digital Content 1, which describes sham AP points in detail).  Participants in the verum and placebo CHM groups were scheduled for clinic visits on weeks 1, 4, 8, and 12 of the treatment period. Women in both groups were instructed to take 3 capsules orally with water twice per day (morning and evening) and to report daily medication intake in their hot flush diary.  Verum CHM Group:  Women received Zhi Mu 14, a standardized CHM preparation comprising 14 plant materials (see Table, Supplemental Digital Content 2, which describes the composition of the CHM formula Zhi Mu 14 in detail). The formulation of Zhi Mu 14 is based on two modified classic formulas VB Gan Mai Da Zao Tang and Qing Hao Bie Jia Tang designed to treat hot flushes, night sweats, insomnia, mood swings, irritability, and emotional instability resulting from kidney yin deficiency. The dose of herbal extract granules (3 g/d) administered is equivalent to 15 g of dry herb.  Placebo CHM Group: women received placebo capsules containing starch (Amylum maydis) and caramel as color tracer. This placebo mixture has no known effects on menopausal symptoms.  Both placebo and CHM capsules were identical in appearance and were prepared by Sheng Foong Pharmaceutical Ltd (Taiwan) on behalf of China Medical Ltd (Aesch, Switzerland). | HF: participants **recorded hot flush severity and frequency per week**, an adaptation of the daily hot flush diary. Hot flush severity score was calculated as the sum of the number of self-reported hot flushes multiplied by severity. The applied hot flush severity rating scale scores were as follows: 1 = mild (heat sensation without sweating and disruption of activity), 2 = moderate (heat sensation accompanied by sweating but with no disruption of activity), and 3 = severe (heat sensation accompanied by sweating and disruption of activity). Participants were asked to record the occurrence and severity of each hot flush during the run-in and treatment periods, and on the 4th, 8th, and 12th weeks of the follow-up period of the trial. | Mood: Menopause related quality of life was assessed using the validated MRS II. This self-report instrument comprises 11 items and assesses the presence and intensity of menopausal symptoms on a 5-pointrating scale ranging from 0 (no symptom) to 4(very severe symptom). A **psychological subscale of MRS II represents the domain specific index of severity of climacteric-related psychological complaints** (ie, 0-4 =none tolittle;5-8=mild;9-16=moderate; 17 =severe). The mean reference value for the MRS II psychological subscale in European populations is 2.9. | HF:  No significant differences in any hot flush mean change value have been found between the verum CHM group and the placebo CHM group (p<90.42; Table 3). | Mood:  In the verum CHM group, MRS II psychological subscale decreased significantly from pretreatment to follow-up assessment only (p<0.020). In the placebo CHM group, there was a significant decrease in the MRS II sum score (P= 0.036),but not in the subscale scores (P’sQ0.08). |

Notes:

MP	Menopausal

PeriMP	Perimenopausal

PostMP	Postmenopausal

VMS	Vasomotor symptoms (includes hot flashes, night sweats)
